# Antivirals for seasonal influenza in the US: modeling direct and indirect effects with variable uptake

**DOI:** 10.64898/2026.09.10.26362674

**Authors:** Sinead E. Morris, Samantha G. Dean, Louis Yat Hin Chan, Shikha Garg, Timothy M. Uyeki, Rebecca K. Borchering, Matthew Biggerstaff, Alexandra M. Mellis

## Abstract

Influenza antivirals can directly benefit treated individuals by reducing the risk of some influenza-associated complications when initiated soon after symptom onset. Antivirals may also reduce onward transmission, and previous modeling studies including such indirect effects have estimated large population-level reductions in influenza-associated disease outcomes. However, these studies often assume an optimistic number of people initiate, and adhere to, prompt treatment. Using a compartmental framework, we modeled population-level reductions in influenza-associated hospitalizations given potential direct and indirect effects of influenza antivirals in the United States. From reported patterns of care-seeking, antiviral prescribing, and treatment adherence, we estimated 1.1–9.7% of symptomatic individuals initiate and adhere to prompt antiviral treatment. We estimated small direct effects of antivirals in example seasons, with 0.9% of hospitalizations prevented when antivirals reduced hospitalization risk by 20%. Assuming antivirals had direct and indirect effects generated slightly larger impacts. For example, up to 3.8% of hospitalizations were prevented when antivirals reduced hospitalization risk and onward transmission by 20%. More substantial impacts (for example, preventing >20% of hospitalizations) were only achieved by increasing initiation and adherence to prompt treatment beyond typically observed levels. Therefore, prompt treatment initiation and adherence are critical factors shaping potential population-level impacts of influenza antivirals.

## Introduction

Seasonal influenza causes an estimated 5–23 million medically attended illnesses and 120,000–710,000 hospitalizations in the United States (US) each year^1^. Influenza antiviral treatment can reduce the duration of illness and the risk of some influenza-associated complications, especially when initiated within 48 hours of symptom onset^2–7^. These direct effects among treated patients may reduce influenza-associated morbidity at the population level^8^. Prompt treatment may also reduce the risk of onward transmission, thereby indirectly protecting uninfected individuals^9,10^.

Several prescription antiviral medications are approved for treatment of influenza in the US. The most commonly prescribed is oseltamivir, which is recommended for persons of all ages and should be taken twice daily for five days^11–13^. Baloxavir marboxil (henceforth referred to as baloxavir) is a newer, single-dose, antiviral that more rapidly suppresses influenza viral load than oseltamivir^14,15^. It is recommended for the treatment of uncomplicated influenza in persons aged ≥5 years who have been symptomatic for less than 48 hours^11^, although uptake has been low since its approval in 2018^13^.

The US Centers for Disease Control and Prevention (CDC) recommends the initiation of antiviral treatment as soon as possible for any individual with suspected or confirmed influenza who is hospitalized; has severe, complicated, or progressive illness; or who is at higher risk for complications^11^. Treatment may also be considered for patients who are not at higher risk if it can be initiated within 48 hours of symptom onset. Despite these recommendations, influenza antiviral prescribing in US clinical settings is often low, even among individuals at higher risk of complications^16–20^. Among those prescribed antivirals, the number who initiate treatment promptly (for example, within 48 hours of symptom onset), and complete the recommended course, is likely even lower due to delays in care-seeking and imperfect adherence to prescriptions^21,22^.

We previously modeled direct effects of influenza antivirals in the US, accounting for reported patterns of care-seeking, prescribing, and treatment adherence, and estimated small overall reductions in influenza-associated hospitalizations^8^. Prior efforts to model antiviral impact whilst including indirect effects have often estimated much larger reductions in influenza-associated morbidity and/or mortality^23–29^. However, such conclusions tend to be associated with optimistic assumptions about the number of individuals receiving prompt treatment compared to recent US data^16–18,30–32^.

Here we model potential direct and indirect effects of influenza antiviral treatment using real world observations to inform our assumptions about the proportion of symptomatic individuals who initiate, and adhere to, prompt treatment. We estimate the reduction in influenza-associated hospitalizations under different antiviral effectiveness scenarios and examine the impact of increasing the proportion of influenza outpatients who undergo prompt antiviral treatment after symptom onset.

## Results

We developed an age-structured ordinary differential equation (ODE) framework to model direct and indirect effects of influenza antiviral treatment among symptomatic outpatients in the US with realistic uptake and adherence assumptions (Figure 1). We assumed antivirals were only effective among symptomatic individuals who sought care and initiated treatment within 48 hours of symptom onset, and who adhered to treatment whilst infectious (henceforth referred to as individuals undergoing ‘prompt treatment’; see Methods for further details). Using plausible ranges for the timing and likelihood of care-seeking for influenza-like illness, antiviral prescribing patterns in outpatient settings, and antiviral treatment adherence, we estimated that the percentage of symptomatic individuals undergoing prompt treatment varied from 1.1–9.7% across age groups (Table 1)^16–22,30–32^.

**Figure 1.**
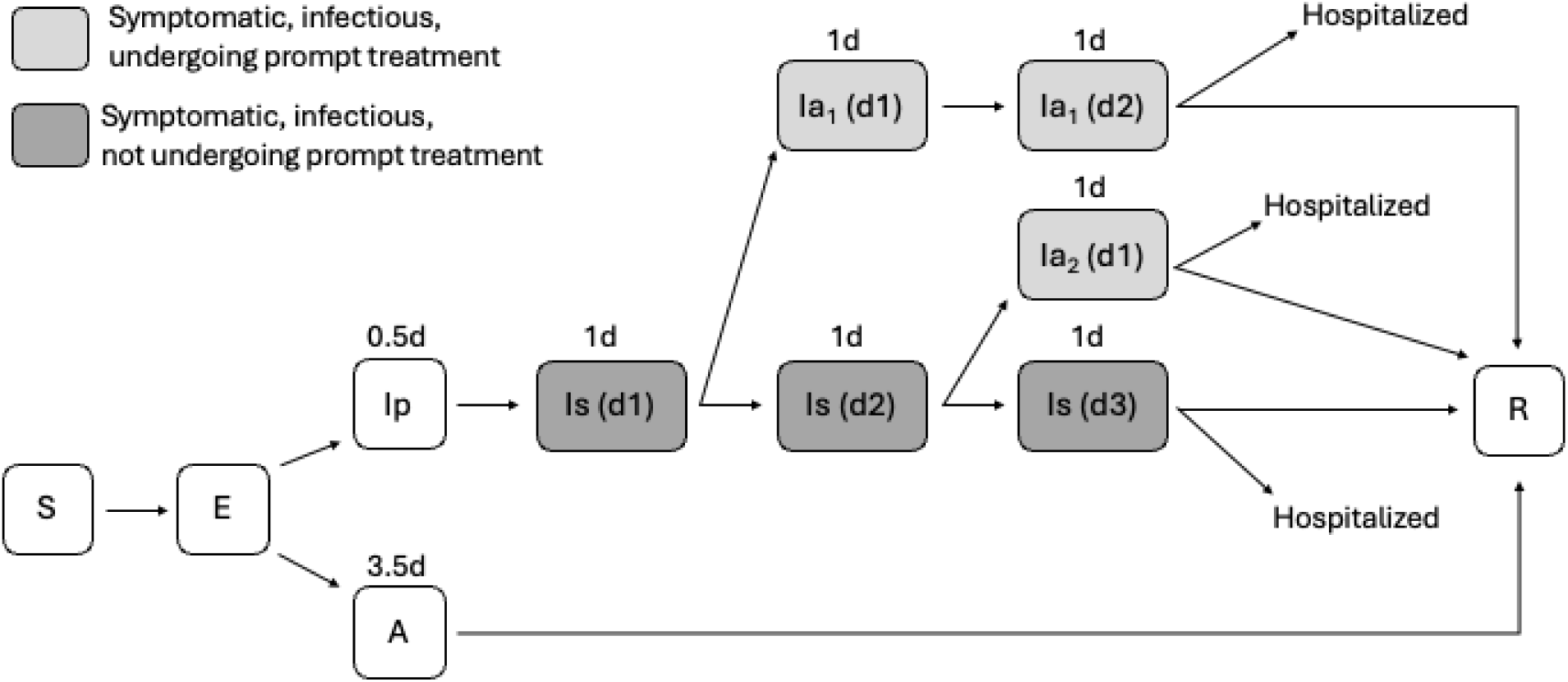
Model structure. (A) Schematic of the ODE model for influenza transmission and antiviral treatment. One age group is shown for clarity; all others follow the same structure. Individuals are either susceptible (S), exposed (E), asymptomatic and infectious (A), pre-symptomatic and infectious (Ip), symptomatic and infectious (Is), symptomatic, infectious, and started antiviral treatment one day after symptom onset (Ia_1_), symptomatic, infectious, and started antiviral treatment two days after symptom onset (Ia_2_), recovered and immune (R), or hospitalized and immune. Numbers above each infectious compartment show its average waiting time. Numbers in parentheses distinguish between symptomatic infectious subcompartments (e.g., ‘Ia1 (d1)’ is the first day (or 0–24 hours) in Ia1 and ‘Ia1 (d2)’ is the second day (or 24–48 hours)). The total average time spent infectious was fixed at 3.5 days for all possible pathways, regardless of symptom or antiviral treatment status. Abbreviations: d, day; ODE, ordinary differential equation.

**Table 1.**
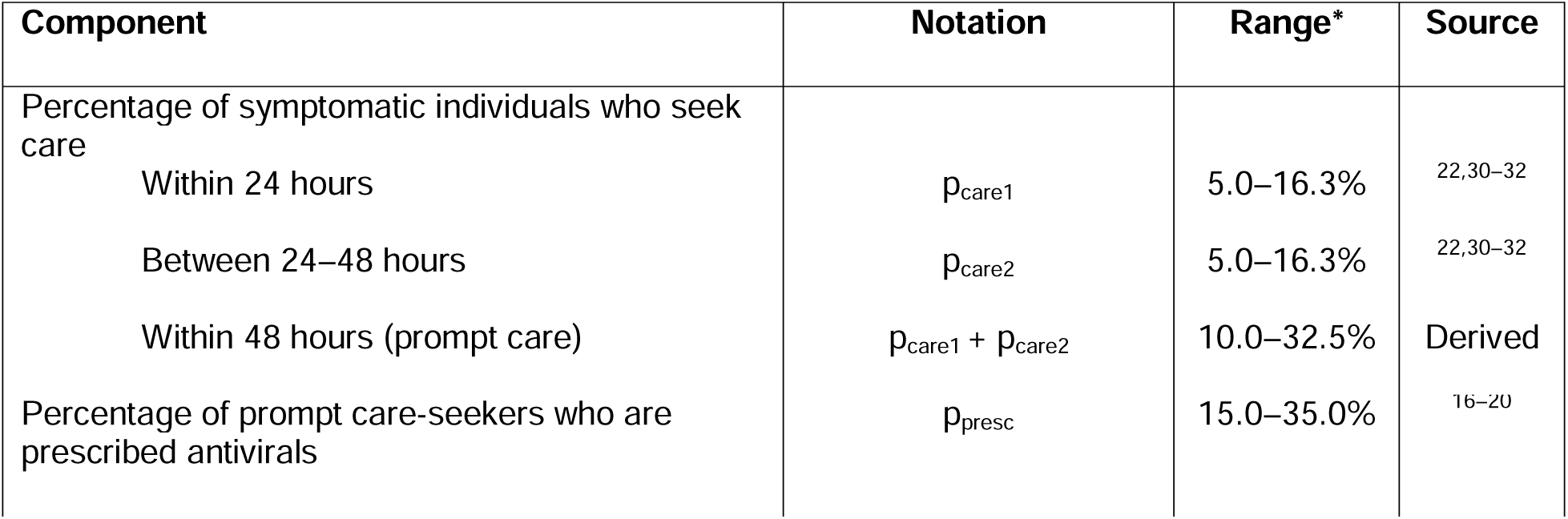
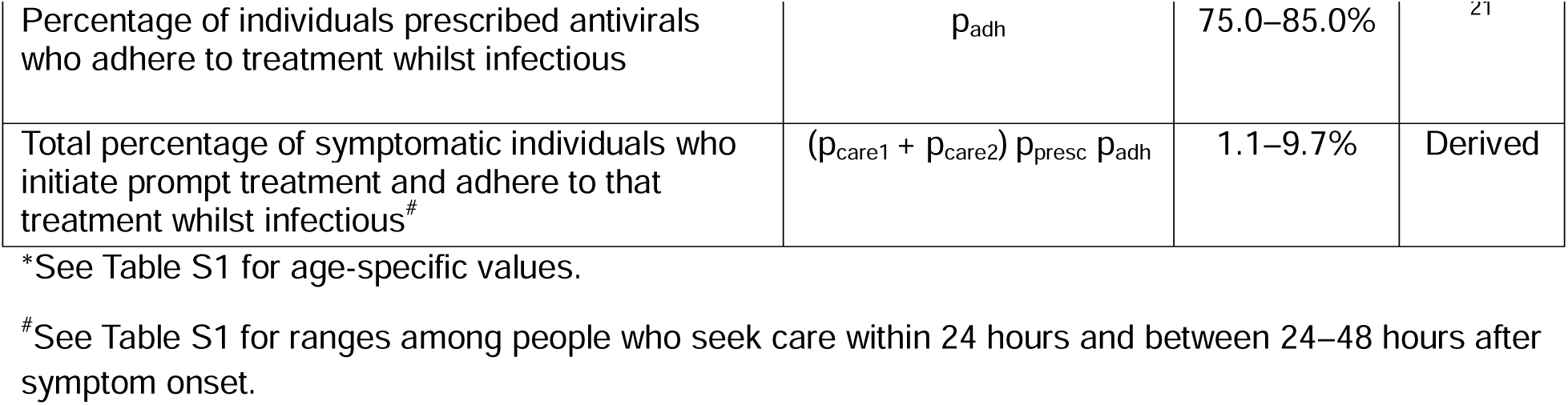
Components determining the percentage of symptomatic individuals who adhere to prompt influenza antiviral treatment. ‘. Prompt care’ refers to care sought within 48 hours of symptom onset and represents the sum of individuals seeking care within 24 hours of symptom onset and those seeking care between 24–48 hours after symptom onset. Uncertainty was incorporated by constructing 100 uniform Latin Hypercube samples from these ranges.

We defined two representative seasons – a low severity season with an initial effective reproduction number (R_e_) equal to 1.1 and a high severity season with R_e_ equal to 1.25 – and explored the impact of antiviral treatment under different assumptions about its effectiveness against influenza-associated hospitalization and onward transmission (Table 2)^2,3,9,10,33–36^. The number of symptomatic illnesses (henceforth referred to as ‘illnesses’) and hospitalizations generated by the model aligned with independent estimates of the number of illnesses and hospitalizations in prior seasons classified as low and high severity by the CDC (Figure 2, Figures S1–S2)^1,37^. The model also reproduced timings of peak illnesses within ranges of historical peaks, and the projected age distribution of illnesses and hospitalizations aligned with previous estimates (Figures S3–S4)^1,38^.

**Figure 2.**
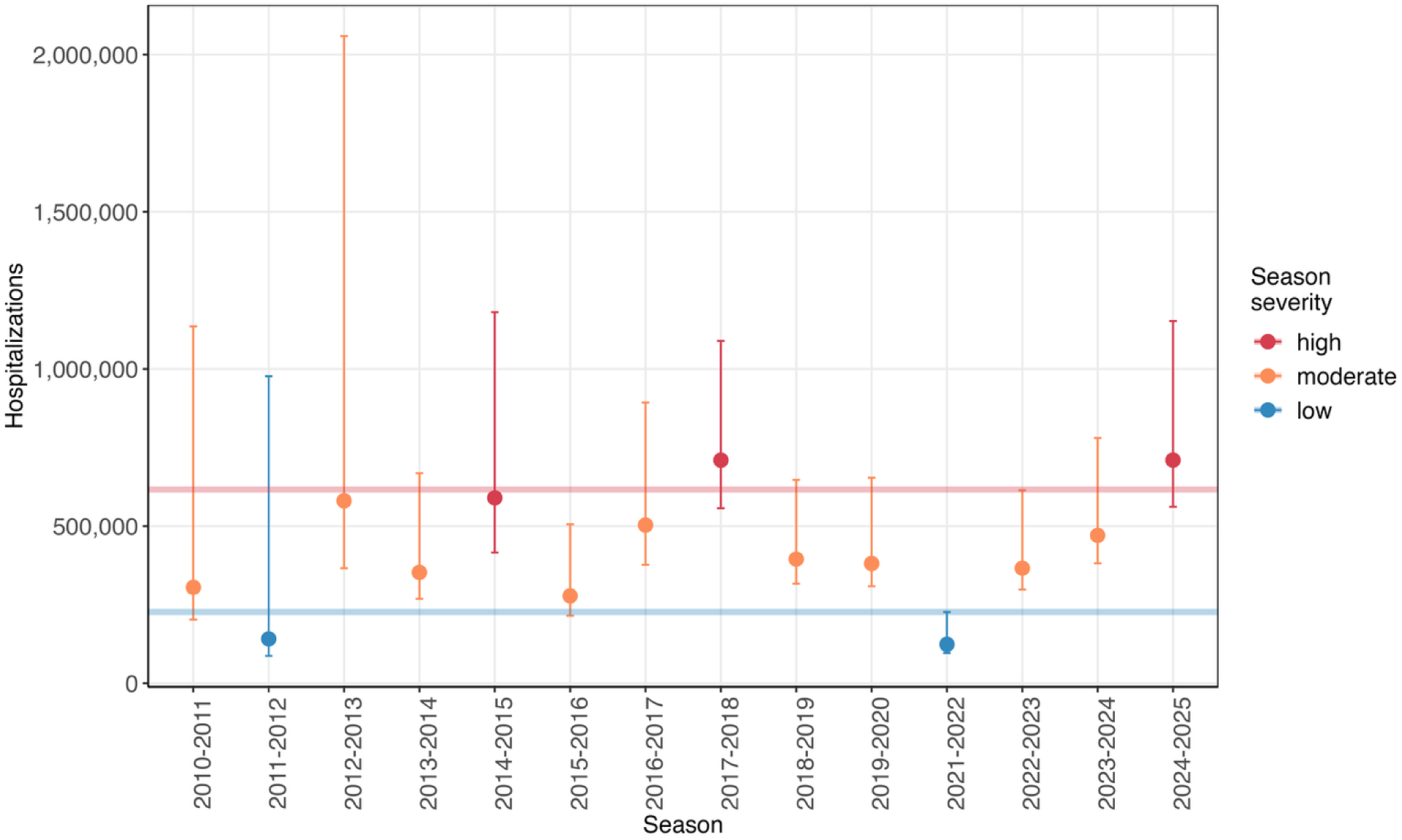
Model validation. Points show independent estimates of the number of influenza-associated hospitalizations in the US in previous influenza seasons and error bars are the 95% uncertainty intervals. Colors show CDC season severity assessments and horizontal lines compare the corresponding mean model outputs for the baseline scenario without antivirals.

**Table 2.** Influenza antiviral effectiveness scenarios.

| Antiviral scenario | Season severity | $R_e$ | Reduction in risk of influenza-associated hospitalization | Reduction in risk of onward transmission |
| --- | --- | --- | --- | --- |
| Baseline (no antivirals) | High<br>Low | 1.25<br>1.10 | –<br>– | –<br>– |
| No transmission impact (direct effects only) | High<br>Low | 1.25<br>1.10 | 20%<br>20% | 0%<br>0% |
| Small transmission impact (direct and indirect effects) | High<br>Low | 1.25<br>1.10 | 20%<br>20% | 10%<br>10% |
| Large transmission impact (direct and indirect effects) | High<br>Low | 1.25<br>1.10 | 20%<br>20% | 20%<br>20% |

\* $R_e$ is the effective reproduction number at the beginning of each simulation.

In the baseline scenario without antivirals, we estimated there would be 227,000 hospitalizations in the low severity season and 616,000 hospitalizations in the high severity season. When antiviral treatment reduced the risk of hospitalization by 20% but did not impact onward transmission (i.e., antivirals had only direct effects), we estimated an average of 2,040 and 5,590 hospitalizations were prevented in the low and high severity seasons, respectively, relative to baseline (Figure 3A). This equated to a 0.9% reduction for both season severities, with 590,000 and 1.50 million people undergoing prompt treatment in the low and high severity seasons, respectively (Table S2). More hospitalizations were prevented when antiviral treatment also had indirect effects, although the percentages were still small relative to baseline. For example, in the large transmission impact scenario (when antivirals reduced the risk of onward transmission by 20%) we estimated a 3.8% and 1.4% reduction in hospitalizations for the low and high severity seasons, respectively, with 573,000 and 1.49 million people undergoing prompt treatment. Although the total number of prevented hospitalizations was greater in the high severity season compared to the low severity season, the converse was true for the number prevented solely by indirect effects (Figure 3B). Similarly, the percent of hospitalizations prevented by indirect effects in the low severity season was always greater than the percent prevented in the high severity season. This is due to the greater relative impact of transmission mitigation when R_e_ is closer to 1. Among age groups, adults ≥65 years experienced the greatest reduction in hospitalizations relative to baseline, although the differences between age groups were minor (Table S3). We also note a small reduction (0.4–0.5%) in total effects across all scenarios when antiviral effectiveness against hospitalization was 10% instead of 20% (Figure S5).

**Figure 3.**
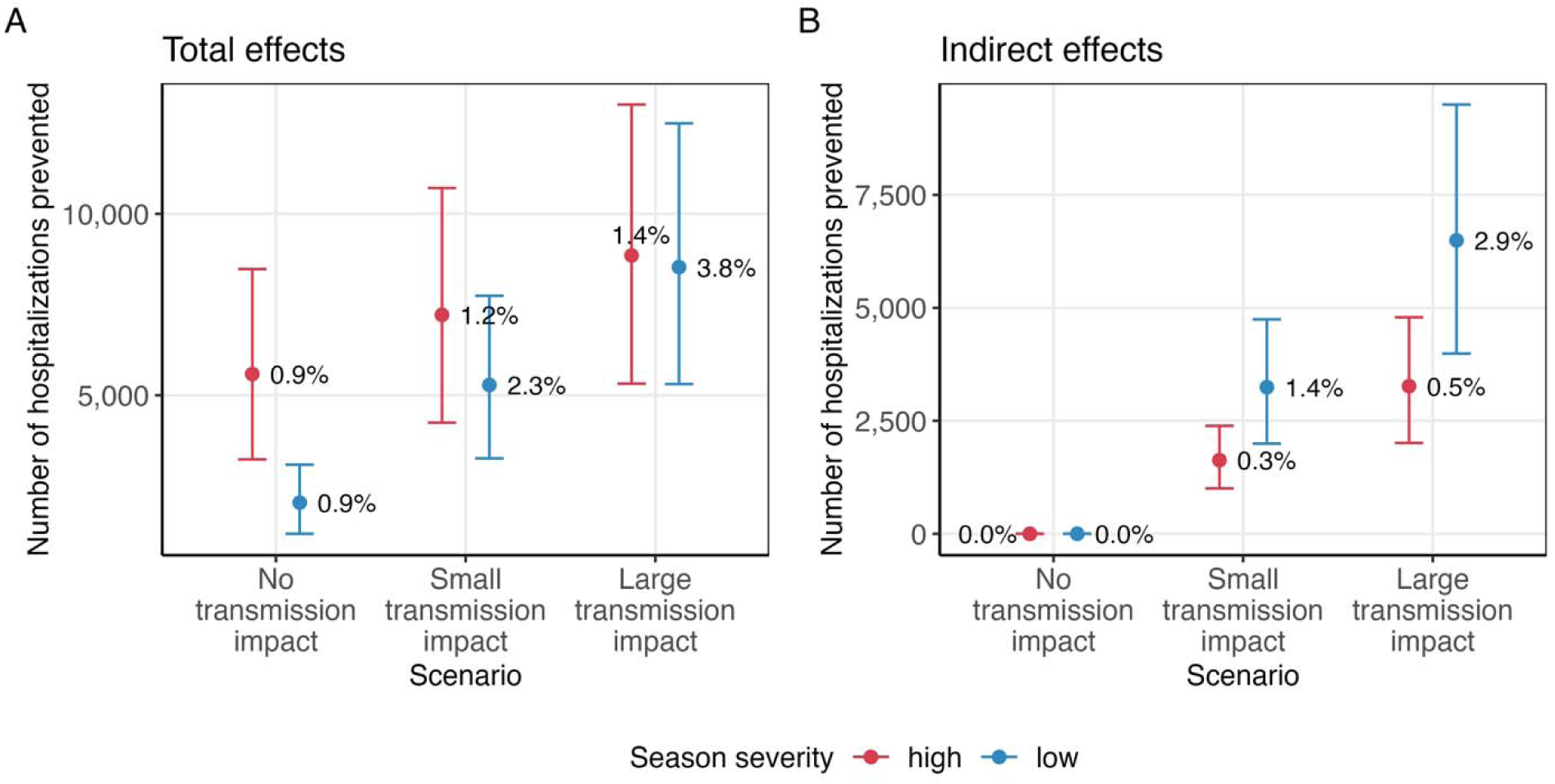
Influenza-associated hospitalizations prevented in three antiviral effectiveness scenarios. (A) Total effects. Points show the mean number of hospitalizations prevented from 100 simulations and error bars are the 95% uncertainty intervals. Text percentages represent the mean percentage of hospitalizations prevented relative to baseline. (B) Same as (A), but for indirect effects. Scenarios are defined in Table 2.

To explore whether low antiviral effectiveness could explain the relatively modest impact of treatment in our original scenarios, we conducted extended simulations in which effectiveness against hospitalization and onward transmission varied from 0–100% (Figure 4A). Even when both effectiveness parameters were set to the unrealistic values of 100%, a maximum of 18.3% and 7.1% of hospitalizations were prevented in the low and high severity seasons, respectively. Antiviral effectiveness against onward transmission had a greater relative impact than antiviral effectiveness against hospitalization. We then varied the percentage of symptomatic individuals undergoing prompt treatment between 0–100%, while fixing antiviral effectiveness against hospitalization at 20%, and estimated much greater potential impacts of antivirals (Figure 4B). For example, increasing the percentage undergoing prompt treatment to 25% (with antiviral effectiveness against onward transmission equal to 20%) prevented 22.5% and 8.0% of hospitalizations in the low and high severity seasons, respectively. Furthermore, up to 99.9% and 96.9% of hospitalizations were prevented when both the percentage undergoing prompt treatment and effectiveness against onward transmission were set to the unrealistically high values of 100% (with effectiveness against hospitalization still fixed at 20%). Results were similar when antiviral effectiveness against hospitalization was fixed at 10%, with up to 99.9% and 96.6% of hospitalizations prevented in the respective seasons (Figure S6). The total number of people undergoing prompt treatment was greatest when assumed antiviral effectiveness against onward transmission was low (Figure S7). This is because weaker indirect effects facilitate increased transmission (and therefore increased burden of illness) in the population.

**Figure 4.**
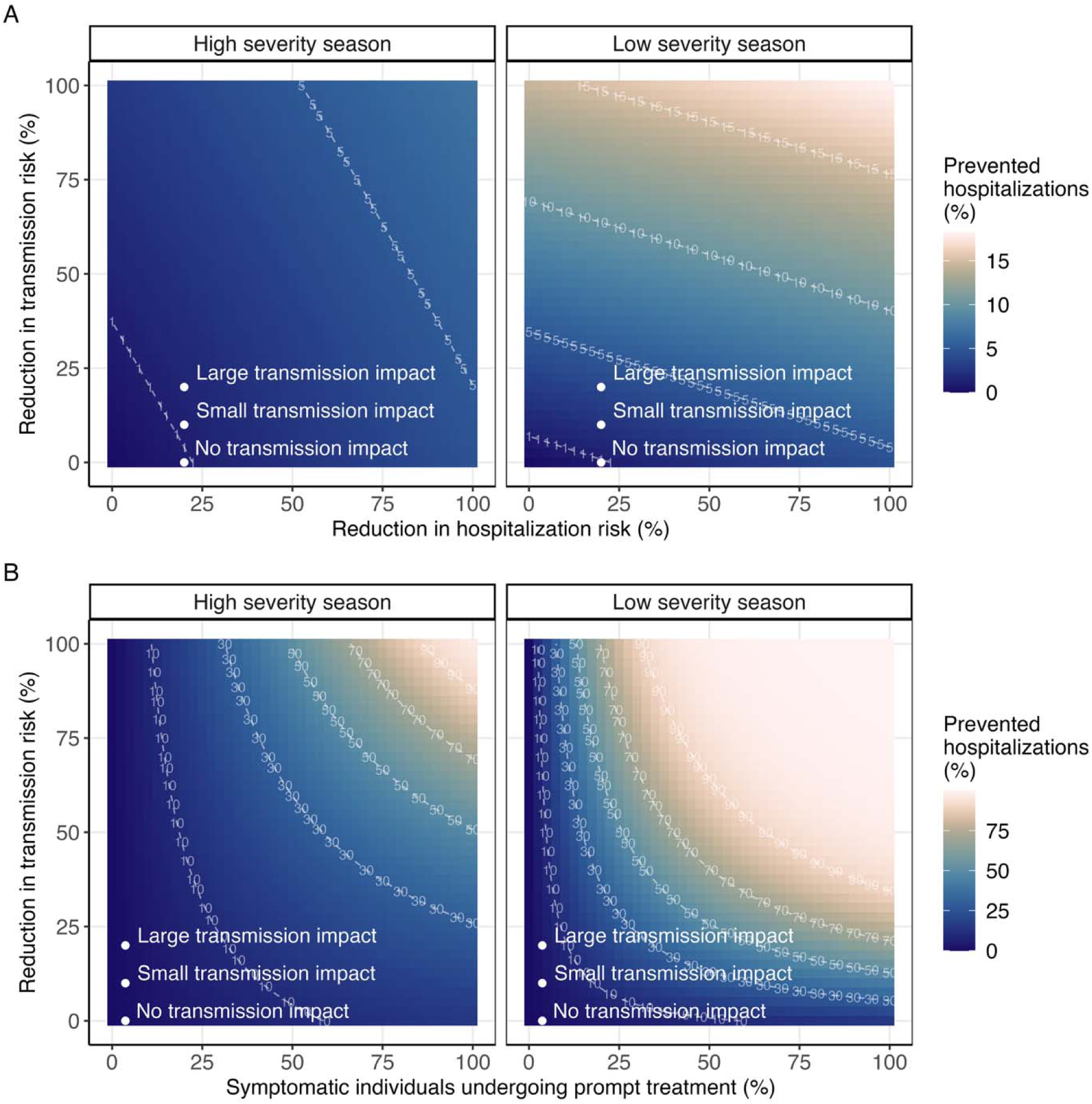
Total percent of influenza-associated hospitalizations prevented across all possible values of antiviral effectiveness and uptake. (A) Percent of hospitalizations prevented relative to baseline for antiviral effectiveness against hospitalization and onward transmission ranging from 0–100%. The percentage of symptomatic individuals undergoing prompt treatment is fixed at its mean value from the original Latin Hypercube samples. Points show the original antiviral effectiveness scenarios. (B) Same as (A), but with antiviral effectiveness against hospitalization fixed at 20%, and the percentage of symptomatic individuals undergoing prompt treatment varying from 0–100%. Note the different color scales in (A) and (B).

In our framework, increases in the percentage of symptomatic individuals undergoing prompt treatment can be achieved through increasing one or more of its underlying components of prompt care-seeking, antiviral prescribing, and adherence (Table 1). We investigated the relative impact of each of these components by returning to our original antiviral effectiveness scenarios (that fixed effectiveness against hospitalization at 20% and effectiveness against transmission at 0%, 10%, or 20%; Table 2). For each scenario and season severity, the biggest singular gain in prevented hospitalizations was obtained when the percentage of symptomatic individuals seeking prompt care (within 48 hours of symptom onset) was increased to 100%, followed by the percentage of prompt care-seekers prescribed antivirals (Figure 5). These were the components with the lowest starting values (Table 1). In contrast, increasing adherence to 100% provided marginal gains. Preventing over 20% of hospitalizations in at least one scenario required increasing two components to 100% (Figure S8). Increasing all three components to 100% (so that all symptomatic infectious individuals underwent prompt treatment) prevented up to 67.4% and 26.1% of hospitalizations in the low and high severity seasons, respectively (Figure S9, large transmission impact scenario).

**Figure 5.**
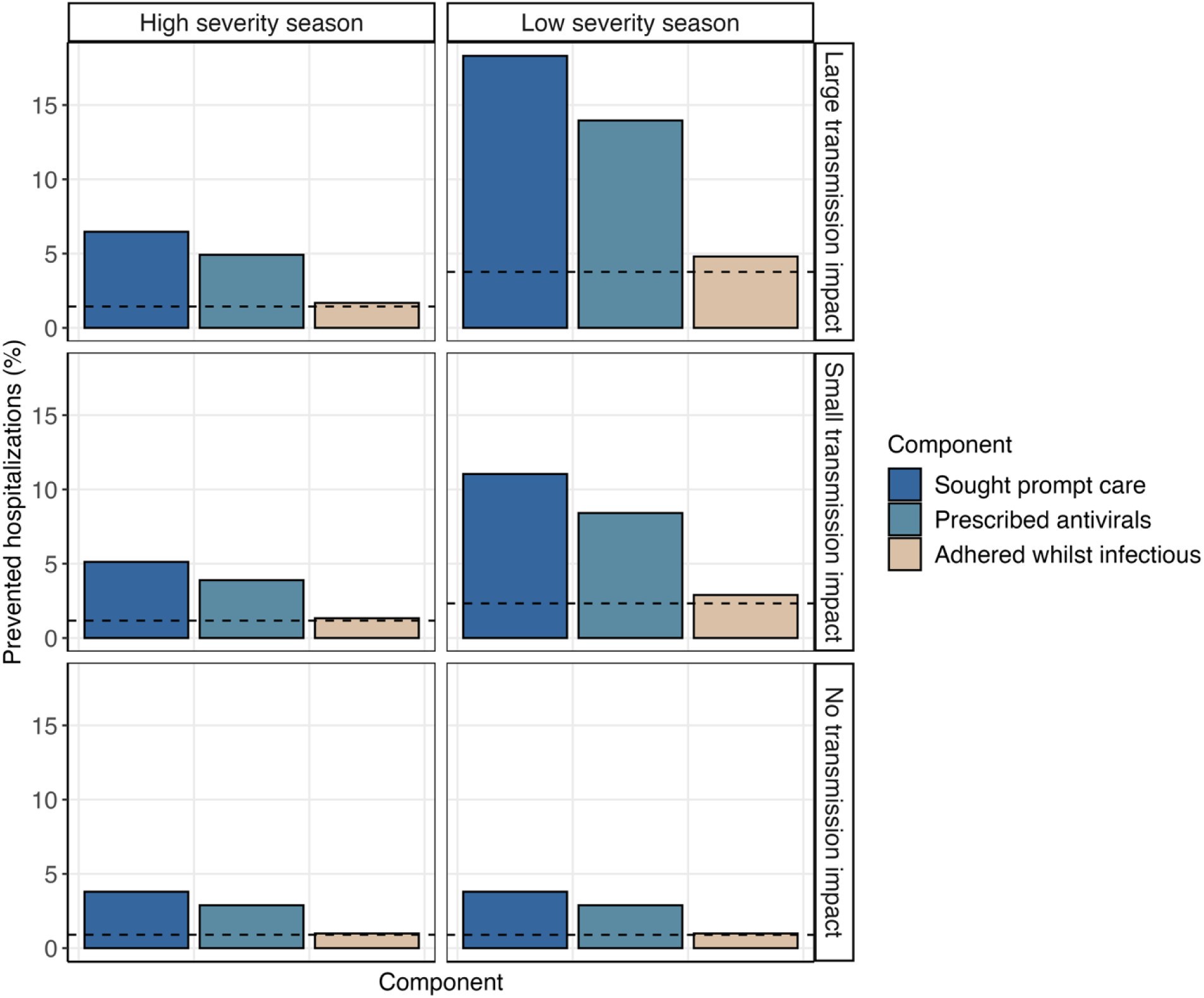
Relative impact of individual components determining the overall percentage of symptomatic individuals undergoing prompt influenza antiviral treatment. Each component was increased to 100% in turn while keeping the other components fixed at their mean starting values. Bars represent the total percent of hospitalizations prevented in each antiviral effectiveness scenario relative to baseline. The dashed horizontal line represents the percent of hospitalizations prevented when all components are fixed at their mean starting values. Antiviral effectiveness scenarios are defined in Table 2. ‘Prompt care’ refers to care sought within 48 hours of symptom onset.

## Discussion

We used an age-structured mathematical model to estimate the impact of influenza antiviral treatment in the US. Under realistic assumptions about antiviral uptake and adherence, we estimated small direct and indirect effects. For example, up to 3.8% of influenza-associated hospitalizations were prevented when antiviral effectiveness against hospitalization and onward transmission were 20%. Even when both parameters were increased to an unrealistically high value of 100%, for illustrative purposes, less than 20% of hospitalizations were prevented at current levels of prompt antiviral treatment. In contrast, increasing the percentage of symptomatic individuals undergoing prompt treatment prevented more hospitalizations than increasing antiviral effectiveness against hospitalization and/or onward transmission. However, multiple improvements in the components influencing this percentage (care-seeking, prescribing, and adherence) were needed to achieve large increases in prevented hospitalizations.

Our estimates of the total impact of antiviral treatment from direct and indirect effects align with one study in the US that assumed similar percentages of individuals undergoing prompt treatment^24^. Other studies that assumed higher likelihoods of prompt treatment have estimated much greater impacts on influenza morbidity and/or mortality. For example, studies with approximately 15–120 million people undergoing prompt treatment have estimated between 25–62% of influenza deaths could be prevented under varying transmission and antiviral effectiveness assumptions^23,25^. Our model can produce similar estimates for prevented hospitalizations if equivalent numbers undergo prompt treatment (Figure 4 and S8). However, achieving such numbers in practice would require large increases in our current best estimates for the percentage of people who initiate and adhere to prompt treatment in the US.

Real-world observations informed our estimates for the likelihood of prompt care-seeking, prescribing, and adherence to influenza antiviral treatment. There are many potential factors underlying the low initiation and adherence suggested by these estimates, including delays to receiving care, under-prescribing, and challenges in accessing and/or adhering to prescribed medications^16,21,22,39^. Reducing barriers to receiving care for influenza illness may enable more responsive care-seeking among some patients. One such example could be through greater use of telemedicine, which has been associated with earlier access to primary care providers compared with in-person visits^40^. Other strategies that could expand access in the community include over-the-counter oseltamivir^41^, although it is not currently approved in the US. In clinical settings, higher levels of prescribing have been reported among patients with a positive influenza laboratory test, and particularly with the use of rapid influenza tests^12,19^.

Therefore, improving access to rapid molecular tests with high sensitivities may increase prescriptions among influenza-positive cases and enable more timely treatment initiation. When appropriate, providers should also be encouraged to follow CDC recommendations for prompt empiric prescribing of influenza antiviral treatment without awaiting laboratory confirmation^11^.

Adherence to treatment may improve with increased use of baloxavir, as only one dose is required. However, uptake has been low since its approval in 2018^13^. We also note that increasing adherence to 100% did not offer substantial improvements in our model unless accompanied by increases in prompt care-seeking and/or prescribing.

Increasing uptake of influenza antivirals may bring additional challenges. First, the actual numbers of antivirals needed may be much higher than the estimates quoted here as most studies do not account for empiric treatment of non-influenza cases (and our study also excluded late treatment of influenza cases and prompt treatment that was discontinued prior to the end of infectiousness). Future work estimating the total number of antivirals needed to treat suspected and confirmed influenza-positive cases would be needed to determine the risks of exceeding availability during periods of high demand^42^. This may be particularly important for pandemic influenza, as our work suggests viruses with higher reproduction numbers (such as those typically estimated for pandemic influenza viruses^33^) will require more people undergoing prompt treatment to achieve similar reductions in the percentage of prevented hospitalizations, all else being equal. Second, greater antiviral uptake may increase the risk of antiviral resistance emerging. Our extended simulations can be used to assess how antiviral impact may be reduced if a resistant strain emerged that impaired its effectiveness. However, they do not address situations in which resistance confers increased transmissibility^43^. Resistance may be more likely to arise against baloxavir than oseltamivir, and continued monitoring of influenza virus susceptibility to antivirals will be important, particularly if there are increases in baloxavir usage^10,25,44,45^.

Our results suggest that influenza antivirals could meaningfully mitigate the population-level burden of influenza-associated hospitalizations, at current assumed levels of direct and indirect effects, if access to prompt treatment can be substantially improved. However, in the absence of major increases in antiviral uptake and adherence, we estimated that the population-level impact will be small. Importantly, our results suggest that increases in antiviral effectiveness alone (for example, through use of different products or improvements in product efficacy^46,47^) are unlikely to substantially reduce influenza-associated hospitalizations in a typical season. We anticipate the impact of antiviral treatment could be even smaller in an influenza pandemic, given that a higher reproduction number was associated with a lower percentage of prevented hospitalizations. Again, our analyses suggest that increased antiviral effectiveness during a pandemic would be unlikely to change this impact unless accompanied by considerable increases in access to prompt treatment.

Compared to antivirals, estimates of the impact of influenza vaccination tend to be higher^48^. For example, in 2022/23 an estimated 24% of influenza-associated hospitalizations were prevented by vaccination: 10% via direct effects and 14% via indirect effects^49^. Although these impacts can vary by season^50^, primarily due to changes in vaccine effectiveness, vaccination can be administered early in an influenza season, prior to infection, and is not subject to challenges of late care-seeking or imperfect adherence. The evidence therefore suggests that influenza vaccination is a more effective way to protect against influenza-associated illness and complications.

There are several limitations to our analysis. First, we assumed that antivirals were not effective if initiated ≥3 days after symptom onset. Although such later treatment is recommended for patients at higher risk for severe influenza or those with complicated or progressive illness, and may reduce the risk of additional complications, it is less likely to impact onward transmission^11,35,36,51,52^. Second, we only required individuals to adhere to treatment whilst infectious to acquire the full benefit of antiviral effectiveness against hospitalization.

Requiring at least five days of treatment would lower our adherence parameter estimate (based on findings from a household study of oseltamivir) and therefore our projected direct effects^21^. However, the overall reduction in prevented hospitalizations may be small as our prior study imposed similar requirements and produced comparable estimates^8^. Third, we assumed the same effectiveness of antivirals initiated 24 and 48 hours after symptom onset. There is clinical evidence that earlier initiation of antivirals (within 24 hours of onset compared to 24–48 hours) accelerates the time to alleviation of symptoms, and modeling studies have also suggested a greater reduction in cumulative risk of onward transmission^4,14,35,53^. Our model captures the latter as individuals who initiate antivirals earlier will experience the effect of transmission mitigation for a longer period. The former would not impact our conclusions unless also accompanied by a reduction in duration of infectiousness, and such a link has yet to be determined. Fourth, hospitalization occurred at the end of the infectious period in our model, consistent with estimates that the median time from symptom onset to hospitalization is three days^35,54,55^. However, any hospitalization that occurs during the infectious period could reduce subsequent transmission due to institution infection prevention and control measures, and therefore lessen the indirect benefits of antivirals^56–59^. Fifth, we did not stratify individuals by the presence or absence of underlying medical conditions associated with increased risk of influenza complications. Although those with increased risk may receive greater direct benefits from antiviral treatment, we expect fewer differences in the mitigation of onward transmission and therefore in the indirect benefits of antivirals^8,60^. Sixth, we did not account for delays from care-seeking to prescription dispensing, or for prescriptions that are never dispensed^12^.

Combined, these gaps could meaningfully reduce the number of people undergoing prompt treatment and lead to even smaller estimated impacts in our original scenario analyses. Finally, we note that care-seeking, prescribing, and dispensing can vary by many factors, including age, race and ethnicity, rurality, clinical setting, influenza testing, and the presence of certain underlying medical conditions^12,19,20,22,61,62^. We model this variation implicitly by including uncertainty in our inputs for the percentage undergoing prompt treatment. However, there may be additional outlying values that we have not considered and our results do not address whether systematic differences in uptake could disproportionately influence antiviral impact among certain groups.

We modeled the impact of influenza antiviral treatment in the US using an age-structured compartmental framework informed by real-world antiviral usage. At realistic uptake and adherence levels for the US, we estimated small direct and indirect effects of antivirals in preventing influenza-associated hospitalizations across a range of effectiveness values.

Achieving meaningful increases in prevented hospitalizations required substantially increasing the percentage of symptomatic individuals initiating and adhering to prompt treatment. Our results suggest that the components influencing this percentage (prompt care-seeking, prescribing, and adherence to treatment) are important factors in improving influenza antiviral impact at the population level.

## Methods

### Mathematical model and baseline parameterization

We developed an age-structured ordinary differential equation (ODE) framework to model the direct and indirect effects of influenza antivirals in the US in a single season (Figure 1).

Susceptible individuals (S) who become infected initially enter an exposed class (E) representing latent infection (individuals who are infected but not yet infectious). Upon becoming infectious, a fraction of individuals enter an asymptomatic and infectious compartment (A) and the remainder enter a pre-symptomatic infectious compartment (Ip), followed by a symptomatically infectious compartment (Is) once their symptoms begin. A proportion of symptomatic individuals enter antiviral treatment compartments 24 hours (Ia_1_) or 48 hours (Ia_2_) after symptom onset and remain there until their infectious period ends. All symptomatic individuals (Is, Ia_1_, and Ia_2_) eventually either recover without hospitalization (R) or are hospitalized, and are considered immune for the remainder of the season (we do not consider reinfection). All asymptomatic individuals eventually recover without hospitalization. The symptomatic compartments are divided into sub-compartments, each with an average duration of one day, to approximate gamma-distributed waiting times. The total average time spent infectious was fixed at 3.5 days for all individuals, regardless of symptom or antiviral treatment status.

We stratified the population into five age groups, 0–4 years, 5–17 years, 18–49 years, 50–64 years, and ≥65 years, and assumed age-specific differences in contact patterns, the probability of developing symptoms, and the risk of hospitalization (Figure S10, Table S4)^1,63–65^. Influenza vaccination in the current season was incorporated implicitly through an additional age-specific reduction in the fraction of individuals developing symptoms. This reduction was calculated as the vaccination coverage among each age group in 2024/25 multiplied by age-specific values of vaccine effectiveness against outpatient illness^66,67^. For the 0–4-year age group, reported coverage among children 6 months–4 years was adjusted to account for the additional population of infants <6 months who are ineligible for vaccination. To account for pre-existing immunity from prior seasons (obtained following vaccination or natural infection), we assumed 70% of the population was susceptible to influenza infection at the beginning of the season^68^. This percentage was fixed across all age groups. Each simulation was run for 365 days, and so we did not model births, deaths, or population ageing. The remaining natural history parameters were informed by prior literature (Table S4)^69–71^.

### Antiviral uptake and effectiveness scenarios

We first defined a baseline scenario in which antivirals were not used and modeled two influenza epidemics that were designed to represent a low severity season (with initial R_e_ = 1.1) and a high severity season (with initial R_e_ = 1.25)^33^. For each season severity, we then explored three additional scenarios with different assumptions about antiviral effectiveness that were informed by prior literature (Table 2). The first scenario assumed that antiviral treatment had only direct effects by incorporating a 20% reduction in the risk of influenza-associated hospitalization but no impact on onward transmission^2,3,34^. We refer to this as the ‘no transmission impact’ scenario. The second and third scenarios introduced indirect effects by assuming a reduced risk of onward transmission, in addition to a reduced risk of hospitalization^9,10,35,36^. In the ‘small transmission impact’ scenario we assumed a 10% reduction in the probability of onward transmission and in the ‘large transmission impact’ scenario we assumed a 20% reduction. To ensure there was only one source of indirect effects, we assumed the same duration of infectiousness for all symptomatic individuals, regardless of treatment status. As a sensitivity analysis, we repeated each scenario assuming antivirals reduced the risk of hospitalization by 10% whilst keeping all effects on transmission the same.

In all simulations, we assumed antivirals were only effective among people who sought care and initiated treatment 24 or 48 hours after symptom onset, and who adhered to treatment whilst infectious (henceforth referred to as individuals undergoing ‘prompt treatment’). We parameterized the probability of a symptomatic individual undergoing prompt treatment using age-specific estimates of the percentage of symptomatic individuals who seek outpatient care within 24 hours or between 24–48 hours after symptom onset, multiplied by the percentage who are prescribed influenza antivirals and who adhere to treatment for the remainder of the infectious period. The latter estimate was based on the fraction of people reporting at least two full days of oseltamivir treatment among people who initiated any oseltamivir treatment in a US household study^21^. Following evidence that most influenza antivirals are prescribed on the day of care-seeking, and most dispensing occurs on the day of prescribing, we did not include an additional delay from care-seeking to treatment initiation^12^. We incorporated uncertainty in the parameters governing the probability of undergoing prompt treatment by constructing 100 uniform Latin Hypercube samples from ranges informed by prior literature (Table 1, Table S1). We then simulated the model for each sample, season severity, and antiviral effectiveness scenario (800 simulations in total).

We extended the scenario analyses described above by conducting simulations in which antiviral effectiveness against hospitalization, antiviral effectiveness against onward transmission, and the percentage of symptomatic individuals undergoing prompt treatment varied from 0–100%. First, we jointly varied antiviral effectiveness against hospitalization and antiviral effectiveness against onward transmission, whilst fixing the percentage of symptomatic individuals undergoing prompt treatment at its mean value from the Latin Hypercube samples for each age group (Table S1). Second, we varied the percentage of symptomatic individuals undergoing prompt treatment and antiviral effectiveness against onward transmission, whilst fixing antiviral effectiveness against hospitalization at 20%. Since our aim was to systematically examine different values for the percentage of symptomatic individuals undergoing prompt treatment, we fixed each value across all age groups. We also assumed the percentages initiating treatment 24 hours and 48 hours after symptom onset were equal. As a sensitivity analysis, we repeated the simulations with antiviral effectiveness against hospitalization fixed at 10% instead of 20%. All other parameters were fixed at their original values.

Finally, we examined the relative impact of increasing each component contributing to the overall percentage of symptomatic individuals undergoing prompt treatment (Table 1). We re-simulated each of our original antiviral effectiveness scenarios while increasing one, two, or all three components to 100% (so that up to 100% of symptomatic individuals underwent prompt treatment). Components that weren’t increased were fixed at their mean values from the original Latin Hypercube samples. When the Latin Hypercube samples varied by age, we took the mean across all age groups. We considered all possible permutations of the three components (e.g., increasing component 1 then 2 then 3 vs. increasing component 2 then 1 then 3), since a component’s impact could differ depending on when in the sequence it was increased.

### Model validation and estimated impacts

We validated the model using independent estimates of illnesses and hospitalizations in the US from 2010–2025, among seasons classified as low or high severity by the CDC^1,37^. We verified that the mean number of illnesses and hospitalizations from our baseline scenario (without antivirals) and our most optimistic antiviral scenario (the large transmission impact scenario) aligned with estimates from these seasons. We also verified that the baseline age distributions of illnesses and hospitalizations matched estimates from the same source, and that peak illnesses occurred within the same time range as historical peaks in the percentage of outpatient visits for influenza-like-illness reported by CDC’s FluView^38^.

We estimated the total effects of antiviral treatment as the difference in hospitalizations between the baseline scenario and each of the three additional antiviral effectiveness scenarios. We express these effects as the absolute number of hospitalizations prevented by antiviral treatment and the percent prevented relative to baseline. We also decomposed the total effects into direct effects and indirect effects as follows. Direct effects were estimated as the difference in hospitalizations between the baseline scenario and the no transmission impact scenario, whereas indirect effects were estimated as the difference between the no transmission impact scenario and the small and large transmission impact scenarios. We present results as the mean and 95% uncertainty interval (2.5^th^–97.5^th^ percentiles) from 100 simulations.

For the extended analyses, we present the percent of hospitalizations prevented relative to the corresponding baseline simulation in which antivirals were not used. We note that with large transmission mitigation impacts (i.e., a large effectiveness against onward transmission or high percetentage of symptomatic individuals undergoing prompt treatment), the epidemic dynamics can be dampened with additional peaks occurring after our 365-day horizon. Since our focus is antiviral impact in a single season, we do not include hospitalizations arising after 365 days when calculating the percent prevented relative to baseline. Although doing so can lead to small decreases in those estimates, our general conclusions are not impacted.

For all analyses, we tracked the number of people undergoing prompt treatment (i.e., the number entering Ia_1_ and Ia_2_) as a measure of antiviral usage, although we do not account for late treatment, prompt treatment that was discontinued prior to the end of infectiousness, or empiric treatment of non-influenza respiratory illness. All analyses were conducted in R version 4.2.2. Absolute numbers are reported to three significant figures and percentages are reported to one decimal place.

## Disclaimer

The findings and conclusions in this report are those of the authors and do not necessarily represent the views of the Centers for Disease Control and Prevention.

## Data availability

All data are publicly available and cited in the text. Code is available at https://github.com/CDCgov/flu-antiviral-ODEmodel.

