## Supplementary Information for "Antivirals for seasonal influenza in the US: modeling direct and indirect effects with variable uptake"

Sinead E. Morris[1,2,*], Samantha G. Dean[1], Louis Yat Hin Chan[1], Shikha Garg[1], Timothy M. Uyeki[1], Rebecca K. Borchering[1], Matthew Biggerstaff[1], Alexandra M. Mellis[1]

[1]Influenza Division, US Centers for Disease Control and Prevention, Atlanta, GA, USA
[2]Goldbelt Professional Services, Chesapeake, VA, USA

#### Model equations

We denote the per-capita transmission between age groups $i$ and $j$ at time $t$ as $\beta_{i,j}(t) = \beta_0 \hat{C}_{i,j}(1 + a\sin(2\pi(t-\phi)/365))$, where $\hat{C}$ is the contact matrix and $(1 + a\sin(2\pi(t-\phi)/365))$ is a seasonal forcing term. For age group $i$,

$$\begin{aligned}
\frac{dS_i}{dt} &= -S_i\Big(\sum_{j=1}^{5}\beta_{i,j}(t)(Is_j + Ip_j + r_{av}Ia_j + r_aA_j)\Big),\\
\frac{dE_i}{dt} &= S_i\Big(\sum_{j=1}^{5}\beta_{i,j}(t)(Is_j + Ip_j + r_{av}Ia_j + r_aA_j)\Big) - \sigma E_i,\\
\frac{dA_i}{dt} &= (1-p_{s,i})\sigma E_i - \gamma_a A_i,\\
\frac{dIp_i}{dt} &= p_{s,i}\sigma E_i - \gamma_p Ip_i,\\
\frac{dIs_{d1,i}}{dt} &= \gamma_p Ip_i - \gamma_s Is_{d1,i},\\
\frac{dIs_{d2,i}}{dt} &= (1-p_{a1,i})\gamma_s Is_{d1,i} - \gamma_s Is_{d2,i},\\
\frac{dIs_{d3,i}}{dt} &= (1-\hat{p}_{a2,i})\gamma_s Is_{d2,i} - \gamma_s Is_{d3,i},\\
\frac{dIa1_{d1,i}}{dt} &= p_{a1,i}\gamma_s Is_{d1,i} - \gamma_s Ia1_{d1,i},\\
\frac{dIa1_{d2,i}}{dt} &= \gamma_s Ia1_{d1,i} - \gamma_s Ia1_{d2,i},\\
\frac{dIa2_{d1,i}}{dt} &= \hat{p}_{a2,i}\gamma_s Is_{d2,i} - \gamma_s Ia2_{d1,i},\\
\frac{dR_i}{dt} &= \gamma_a A_i + \gamma_s Is_{d3,i} + \gamma_s Ia1_{d2,i} + \gamma_s Ia2_{d1,i}.
\end{aligned}$$

Here, $\hat{p}_{a2,i} = p_{a2,i}/(1-p_{a1,i})$ is the adjusted proportion entering $Ia2_{d1,i}$ after accounting for those already entering $Ia1_{d1,i}$. The sums of the different symptomatic infectious compartments are written as $Is_i = Is_{d1,i} + Is_{d2,i} + Is_{d3,i}$, and $Ia_i = Ia1_{d1,i} + Ia1_{d2,i} + Ia2_{d1,i}$. We determine the number of hospitalizations as a proportion of those recovering from symptomatic infection: $h_i\gamma_s(Is_{d3,i} + r_{h,i}(Ia1_{d2,i} + Ia2_{d1,i}))$. Variables are defined in the main text (see also Figure 1) and parameters are defined in Table S4.

#### Effective reproduction number

We derived an expression for the effective reproduction number in the absence of antiviral treatment (i.e., $p_{a1,i} = p_{a2,i} = 0$). First, we approximated the contribution, $C$, of each infectious compartment to the overall reproduction number as the probability an infectious individual would enter the compartment multiplied by the average duration spent in the compartment. This gave the following expressions for an infectious individual from age group $i$,

$$
\begin{aligned}
C_{A,i} &= (1 - p_{s,i})/\gamma_a, \\
C_{Ip,i} &= p_{s,i}/\gamma_p, \\
C_{Is,i} &= p_{s,i}(3/\gamma_s),
\end{aligned}
$$

where $C_{Is,i} = C_{Is_{d1},i} + C_{Is_{d2},i} + C_{Is_{d3},i}$. We then defined the effective reproduction number as

$$R_e = \beta_0(1 - p_R)E(K),$$

where $p_R$ is the proportion of the population immune at the beginning of the simulation and E(K) is the maximum eigenvalue of the matrix K with entries

$$K_{i,j} = \hat{C}_{i,j} N_i (r_a C_{A,j} + C_{Ip,j} + C_{Is,j}).$$

#### Supplementary tables

**Table S1 – Parameterization of the percentage of symptomatic individuals undergoing prompt influenza antiviral treatment, including age-specific ranges.**

| Parameter | Notation | Age | Range | Source(s) |
|---|---|---|---|---|
| Percentage of symptomatic infectious individuals who seek care (at any time) | $p_{any}$ | 0–4y<br>5–17y<br>18–49y<br>50–64y<br>$\geq$ 65y | 25.0–35.0%<br>25.0–35.0%<br>35.0–50.0%<br>40.0–55.0%<br>45.0–65.0% | [1] |
| Percentage of care-seekers who seek care within 24 hours of symptom onset | $p_{c1}$ | | 20.0–25.0% | [2-4] |
| Percentage of care-seekers who seek care 24–48 hours after symptom onset | $p_{c2}$ | | 20.0–25.0% | [2-4] |
| Percentage of symptomatic infectious individuals who seek care within 24 hours of symptom onset | $p_{care1} = p_{any} \times p_{c1}$ | 0–4y<br>5–17y<br>18–49y<br>50–64y<br>$\geq$ 65y | 5.0–8.8%<br>5.0–8.8%<br>7.0–12.5%<br>8.0–13.8%<br>9.0–16.3% | Derived |
| Percentage of symptomatic infectious individuals who seek care 24–48 hours after symptom onset | $p_{care2} = p_{any} \times p_{c2}$ | 0–4y<br>5–17y<br>18–49y<br>50–64y<br>$\geq$ 65y | 5.0–8.8%<br>5.0–8.8%<br>7.0–12.5%<br>8.0–13.8%<br>9.0–16.3% | Derived |
| Percentage of prompt care-seekers who are prescribed antivirals | $p_{presc}$ | | 15.0–35.0% | [5-9] |
| Percentage of individuals prescribed antivirals who adhere to treatment for $\geq$2 days | $p_{adh}$ | | 75.0–85.0% | [10] |
| Percentage of symptomatic infectious individuals who seek care within 24 hours of symptom onset and complete $\geq$2 days of treatment | $p_{a1} = p_{care1} \times p_{presc} \times p_{adh}$ | 0–4y<br>5–17y<br>18–49y<br>50–64y<br>$\geq$ 65y | 0.6–2.6%<br>0.6–2.6%<br>0.8–3.7%<br>0.9–4.1%<br>1.0–4.8% | Derived |
| Percentage of symptomatic infectious individuals who seek care 24–48 hours after symptom onset and complete $\geq$2 days of treatment | $p_{a2} = p_{care2} \times p_{presc} \times p_{adh}$ | 0–4y<br>5–17y<br>18–49y<br>50–64y<br>$\geq$ 65y | 0.6–2.6%<br>0.6–2.6%<br>0.8–3.7%<br>0.9–4.1%<br>1.0–4.8% | Derived |

**Table S2 – Number of people undergoing prompt influenza antiviral treatment.** In these scenarios, antiviral effectiveness against hospitalization is fixed at 20%.

| Antiviral effectiveness scenario | High severity season | Low severity season |
|---|---|---|
| No transmission impact | 1,500,000 (927,000–2,150,000) | 590,000 (365,000–847,000) |
| Small transmission impact | 1,500,000 (925,000–2,150,000) | 582,000 (361,000–830,000) |
| Large transmission impact | 1,490,000 (924,000–2,140,000) | 573,000 (358,000–812,000) |

**Table S3 – Total percent of influenza-associated hospitalizations prevented by age group in each antiviral effectiveness scenario relative to the baseline scenario without antivirals.**

| | No transmission impact | | Small transmission impact | | Large transmission impact | |
|---|---|---|---|---|---|---|
| | % prevented within group | % of all prevented | % prevented within group | % of all prevented | % prevented within group | % of all prevented |
| High severity season | | | | | | |
| 0-4 yrs | 0.5 | 1.9 | 0.8 | 2.2 | 1.0 | 2.3 |
| 5-17 yrs | 0.5 | 2.3 | 0.7 | 2.4 | 0.9 | 2.5 |
| 18-49 yrs | 0.8 | 13.3 | 1.0 | 13.2 | 1.2 | 13.1 |
| 50-64 yrs | 0.9 | 12.2 | 1.1 | 12.1 | 1.3 | 12.0 |
| ≥65 yrs | 1.0 | 70.3 | 1.3 | 70.1 | 1.6 | 70.0 |
| **Total** | 0.9 | 100.0 | 1.2 | 100.0 | 1.4 | 100.0 |
| Low severity season | | | | | | |
| 0-4 yrs | 0.5 | 2.0 | 2.0 | 2.8 | 3.4 | 3.0 |
| 5-17 yrs | 0.5 | 2.6 | 1.9 | 3.4 | 3.2 | 3.6 |
| 18-49 yrs | 0.8 | 14.7 | 2.1 | 15.7 | 3.5 | 15.9 |
| 50-64 yrs | 0.9 | 13.0 | 2.2 | 13.1 | 3.6 | 13.2 |
| ≥65 yrs | 1.0 | 67.7 | 2.5 | 65.0 | 3.9 | 64.3 |
| **Total** | 0.9 | 100.0 | 2.3 | 100.0 | 3.8 | 100.0 |

% prevented within group: number prevented in age group / baseline number in age group.
% of all prevented: number prevented in age group / total number prevented.

**Table S4 – Input parameter values.** Index $i$ refers to age group, y refers to years.

| Parameter | Notation | Age | Value | Source |
|---|---|---|---|---|
| Population size* | $N_i$ | 0–4y | 18.6 million | [11] |
| | | 5–17y | 54.5 million | |
| | | 18–49y | 143.7 million | |
| | | 50–64y | 62.1 million | |
| | | ≥65y | 61.2 million | |
| Effective reproduction number | $R_e$ | | See Table 2 | [12] |
| Baseline transmission | $\beta_0$ | | Derived from $R_e$ | |
| Contact matrix | $\hat{C}$ | | See Figure S10 | [13] |
| Amplitude of seasonal forcing | $a$ | | 0.1 | Calibrated |
| Phase of seasonal forcing | $\phi$ | | 45 days | Calibrated |
| Average duration of latent infection | $1/\sigma$ | | 1.5 days | [14] |
| Average duration of pre-symptomatic infection | $1/\gamma_p$ | | 0.5 days | [15] |
| Average duration of symptomatic infection (regardless of antiviral treatment) | $3/\gamma_s$ | | 3 days | [15] |
| Average duration of asymptomatic infection | $1/\gamma_a$ | | 3.5 days | Assumption |
| Relative transmissibility of asymptomatic infection | $r_a$ | | 0.6 | [16] |
| Risk of hospitalization (in the absence of antiviral treatment) | $h_i$ | 0–4y | 0.007 | [17] |
| | | 5–17y | 0.003 | |
| | | 18–49y | 0.006 | |
| | | 50–64y | 0.01 | |
| | | ≥65y | 0.09 | |
| Proportion initially immune | $p_R$ | | 0.3 | [18] |
| Vaccination coverage | $v_{c_i}$ | 0–4y** | 0.46 | [19] |
| | | 5–17y | 0.48 | |
| | | 18–49y | 0.31 | |
| | | 50–64y | 0.42 | |

| Parameter | Notation | Age | Value | Source |
|---|---|---|---|---|
| | | $\geq$ 65y | 0.63 | |
| Vaccine effectiveness against developing symptoms | $v_{e_i}$ | 0–4y | 0.50 | [20] |
| | | 5–17y | 0.45 | |
| | | 18–49y | 0.40 | |
| | | 50–64y | 0.45 | |
| | | $\geq$ 65y | 0.40 | |
| Proportion infected who develop symptoms (in the absence of vaccination) | $p_{s_0,i}$ | 0–4y | 0.80 | [21] |
| | | 5–17y | 0.55 | |
| | | 18–49y | 0.40 | |
| | | 50–64y | 0.50 | |
| | | $\geq$ 65y | 0.55 | |
| Proportion infected who develop symptoms (in the presence of vaccination) | $p_{s,i}$ | | $p_{s_0,i}(1 - v_{c_i} v_{e_i})$ | |
| Reduction in risk of hospitalization following prompt antiviral treatment | $1 - r_{h,i}$ | | See Table 2 | |
| Reduction in transmissibility of infection while undergoing prompt antiviral treatment | $1 - r_{av}$ | | See Table 2 | |
| Proportion undergoing prompt antiviral treatment (1 day after symptom onset) | $p_{a_1,i}$ | | See Table S1 | |
| Proportion undergoing prompt antiviral treatment (2 days after symptom onset) | $p_{a_2,i}$ | | See Table S1 | |

*Population of 340 million with 6%, 16%, 42%, 18%, and 18% in the 0–4, 5–17, 18–49, 50–64 and $\geq$65 year age groups, respectively.

**Reported vaccination coverage among children 6 months–4 years (0.57) was adjusted to account for the additional population of infants <6 months who are ineligible for vaccination.

### Supplementary figures

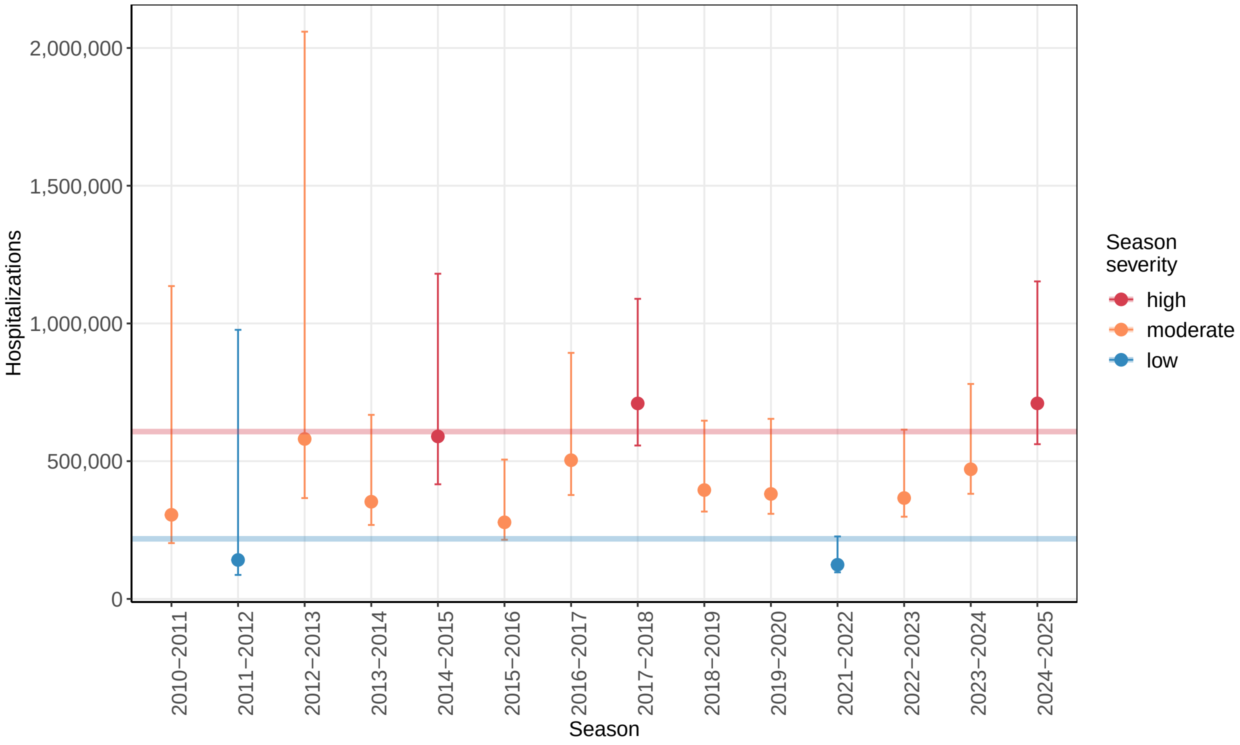


**Figure S1 – Model validation with influenza-associated hospitalizations in the large transmission impact scenario.** Points show independent estimates of the total number of influenza-associated hospitalizations in the US in previous influenza seasons and error bars are the 95th percentile uncertainty intervals. Colors show CDC season severity assessments and horizontal lines compare the corresponding mean model outputs for the most optimistic antiviral effectiveness scenario (large transmission impact).

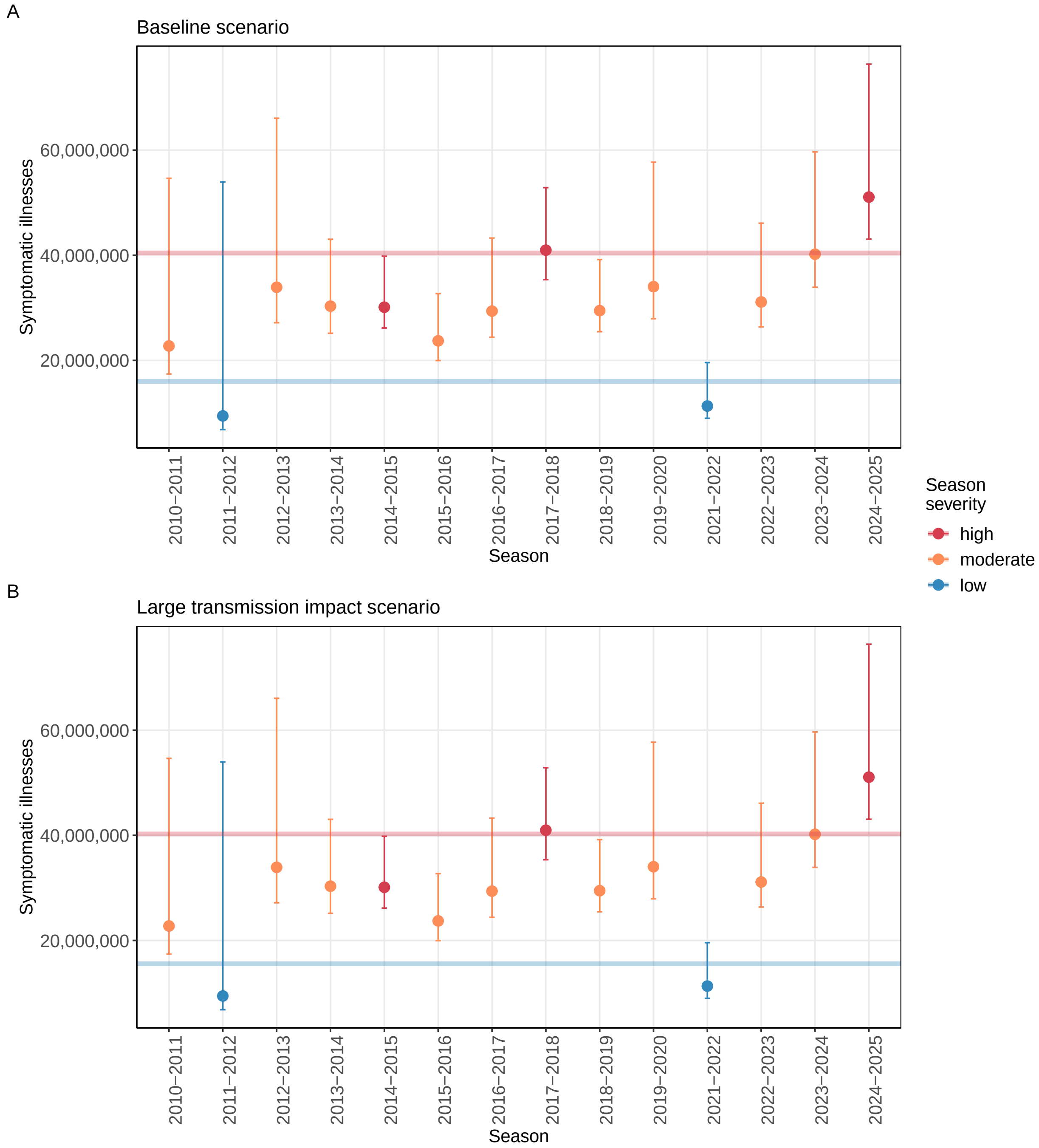


**Figure S2 – Model validation with influenza-associated symptomatic illnesses.** Points show independent estimates of the total number of influenza-associated symptomatic illnesses in the US in previous influenza seasons and error bars are the 95th percentile uncertainty intervals. Colors show CDC season severity assessments and horizontal lines compare the corresponding mean model outputs for the baseline scenario (A) or the most optimistic antiviral effectiveness scenario (large transmission impact) (B). The baseline and large transmission impact scenario outputs appear similar at this scale.

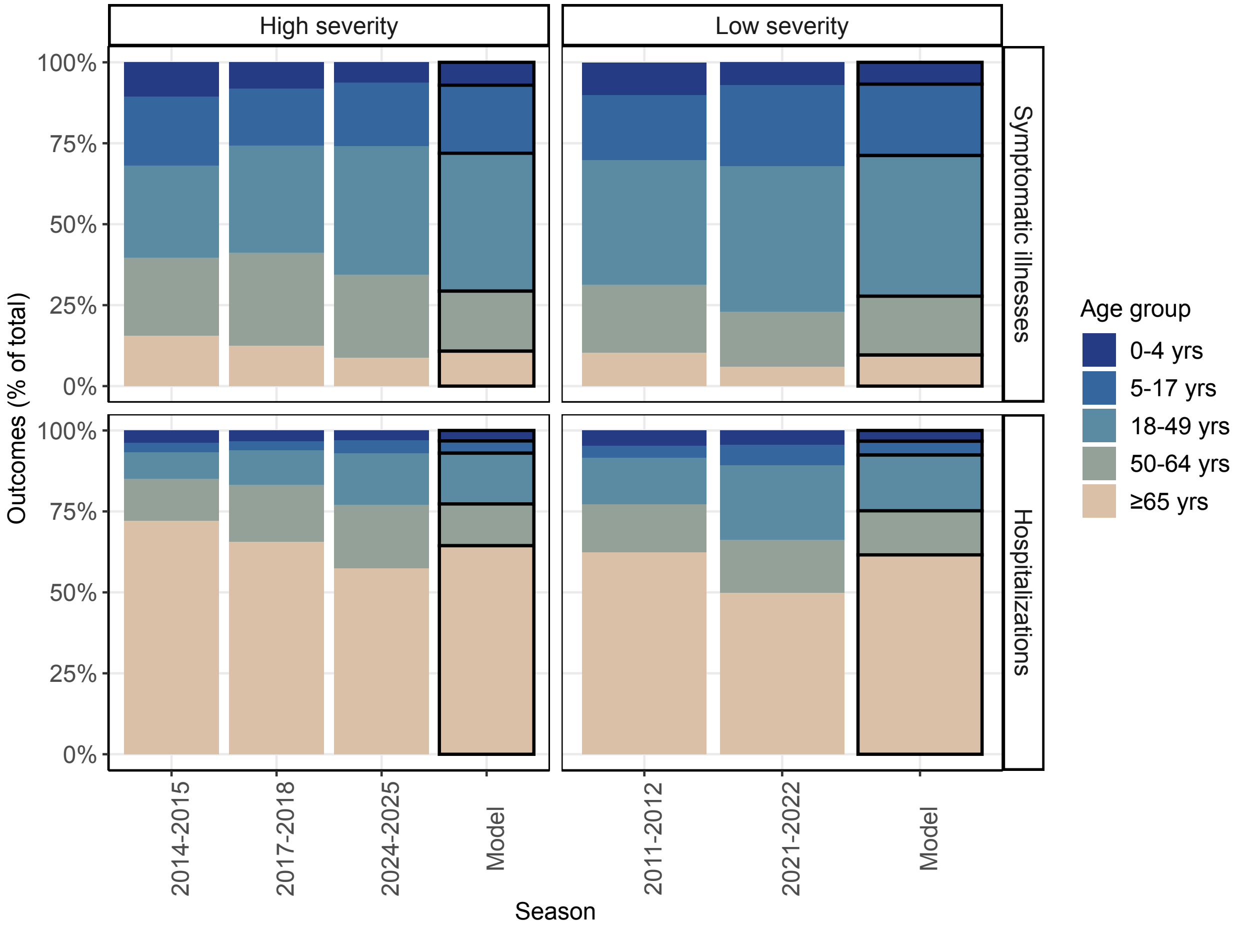


**Figure S3 – Model validation with age distributions** Percentages of each influenza-associated outcome (top – symptomatic illnesses, bottom – hospitalizations) occurring among each age group. Independent estimates from prior seasons are organized by season severity and the corresponding model output is presented alongside and outlined in black.

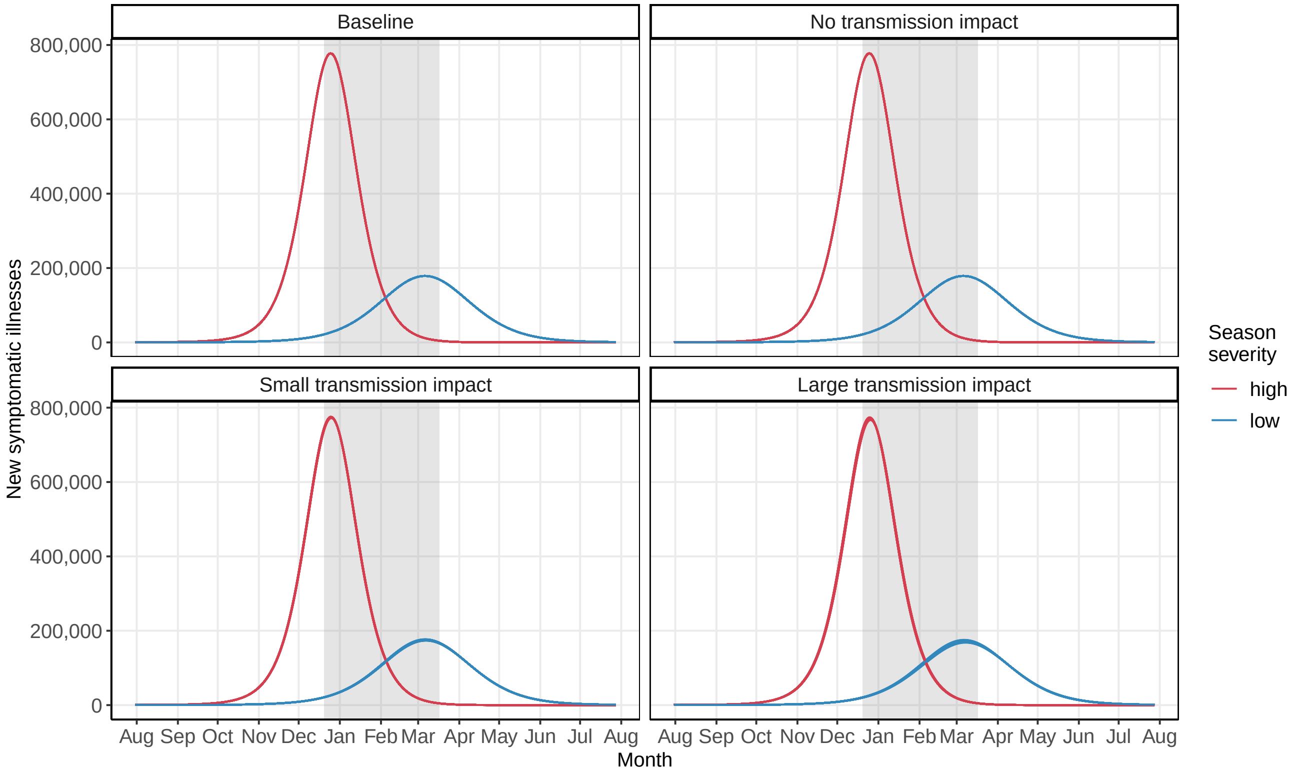


**Figure S4 – Model validation with peak timing.** Lines depict new influenza-associated symptomatic illnesses over time for each of the 100 original Latin Hypercube samples. Grey shaded regions show when the percent of outpatient visits for influenza-like illness has peaked in prior seasons from 2010/11–2024/25 (excluding the 2022/23 season which was atypically early).

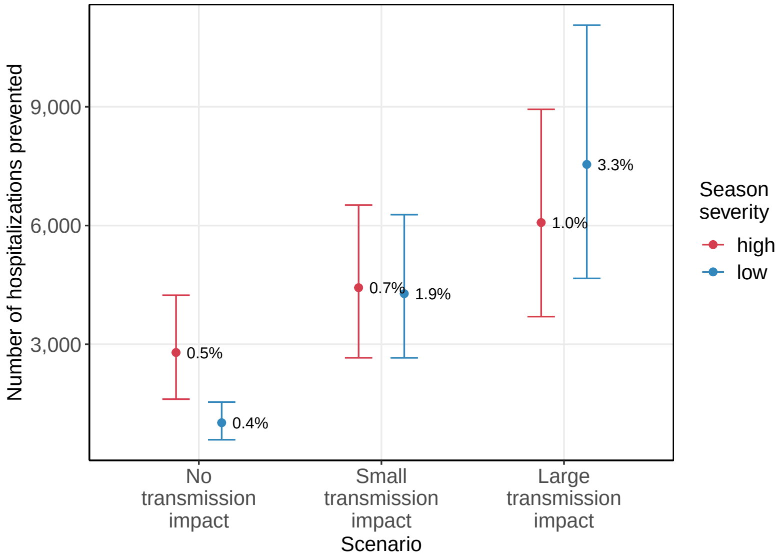


**Figure S5 – Influenza-associated hospitalizations prevented in three antiviral effectiveness scenarios when antiviral effectiveness against hospitalization is 10%.** Points show the mean number of hospitalizations prevented from 100 simulations and error bars are the 95% uncertainty intervals. Text percentages represent the mean percentage of hospitalizations prevented relative to baseline.

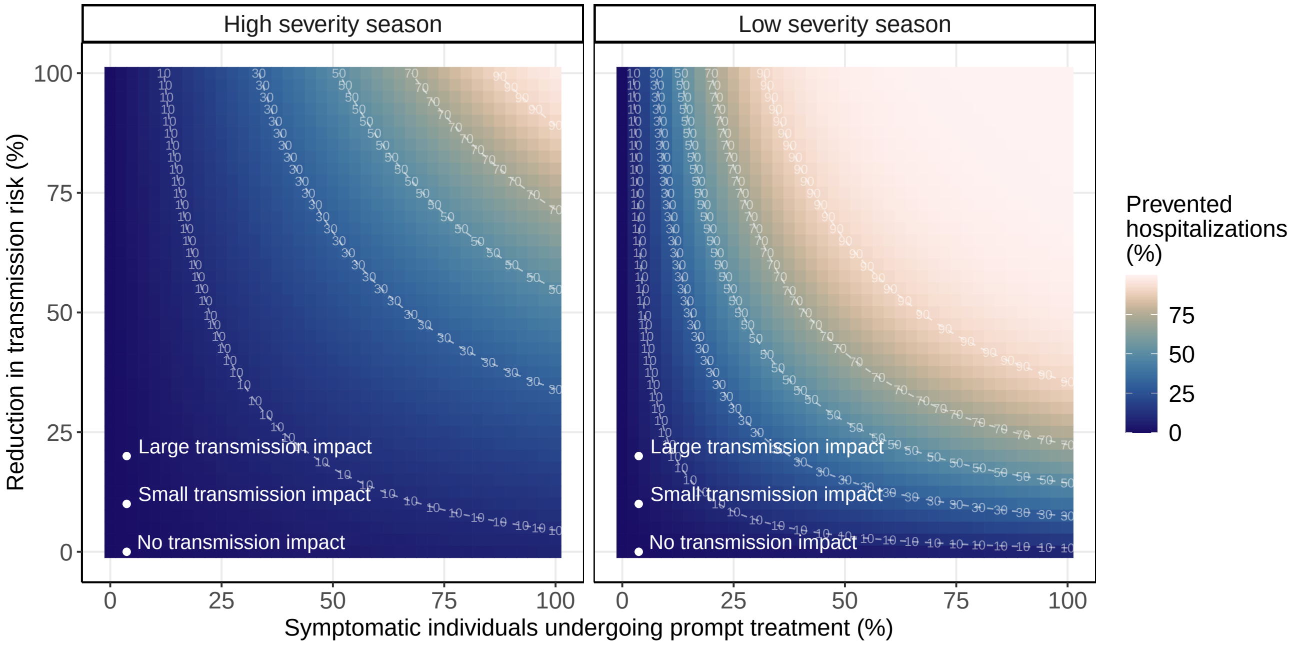


**Figure S6 – Total percent of influenza-associated hospitalizations prevented when antiviral effectiveness against hospitalization is fixed at 10%.** Antiviral effectiveness against onward transmission and the percentage of symptomatic individuals undergoing prompt treatment range from 0–100%. Points show the original antiviral effectiveness scenarios.

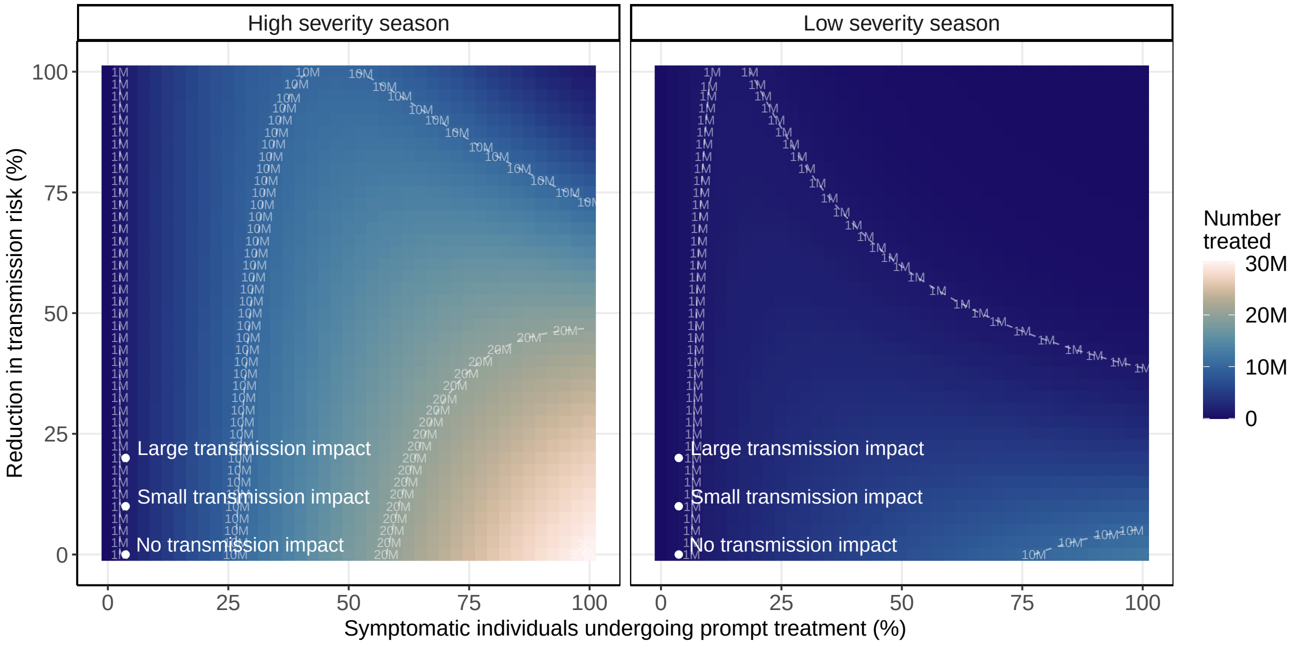


**Figure S7 – Number of individuals undergoing prompt influenza antiviral treatment.** Antiviral effectiveness against onward transmission and the percentage of symptomatic individuals undergoing prompt treatment range from 0–100%. Antiviral effectiveness against hospitalization is fixed at 20%. Points show the original antiviral effectiveness scenarios.

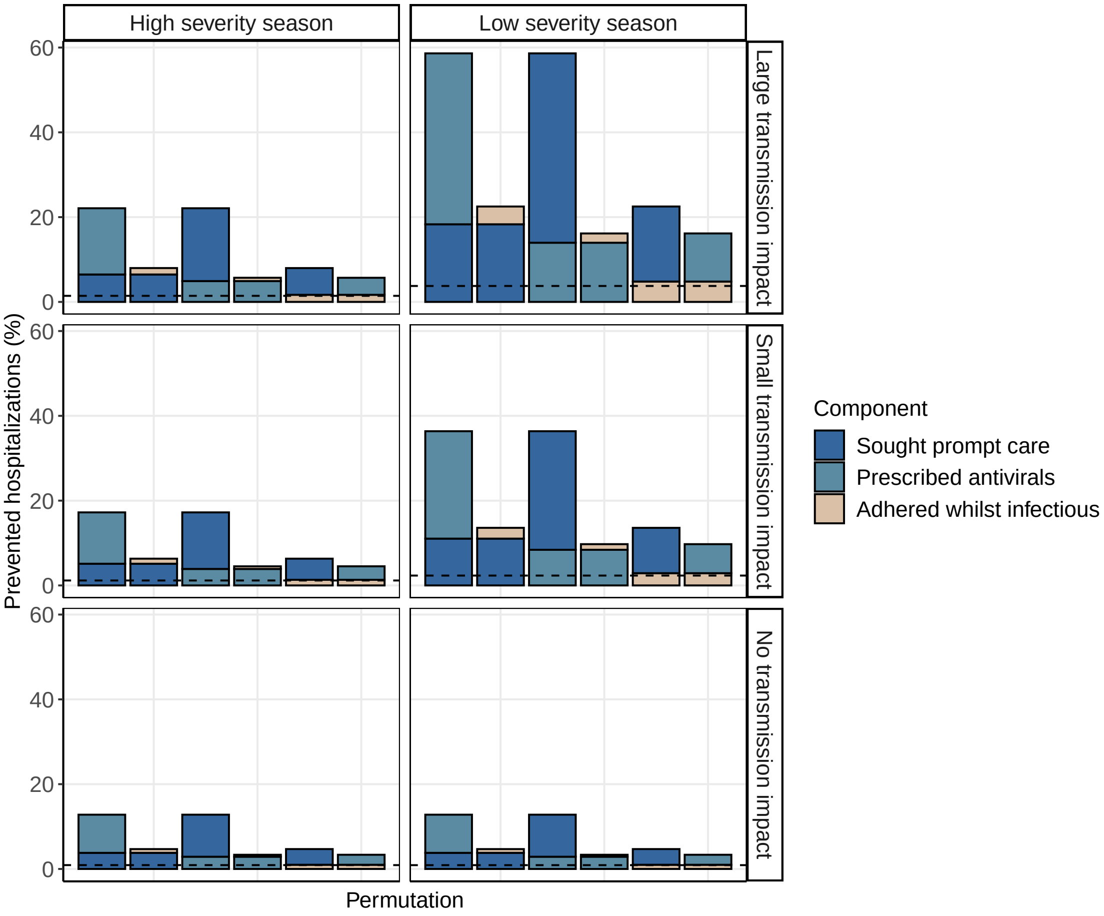


**Figure S8 – Relative impact of increasing two components that determine the overall percentage of symptomatic individuals undergoing prompt influenza antiviral treatment.** Two components were increased to 100% in turn while keeping the other component fixed at its mean starting value. Permutation refers to the different possible orders in which components can be increased; colors at the base of each bar indicate the component that was increased first. Bars represent the total percent of influenza-associated hospitalizations prevented in each antiviral effectiveness scenario relative to baseline. The dashed horizontal line represents the percent of hospitalizations prevented when all components are fixed at their mean starting values. ‘Prompt care’ refers to care sought within 48 hours of symptom onset.

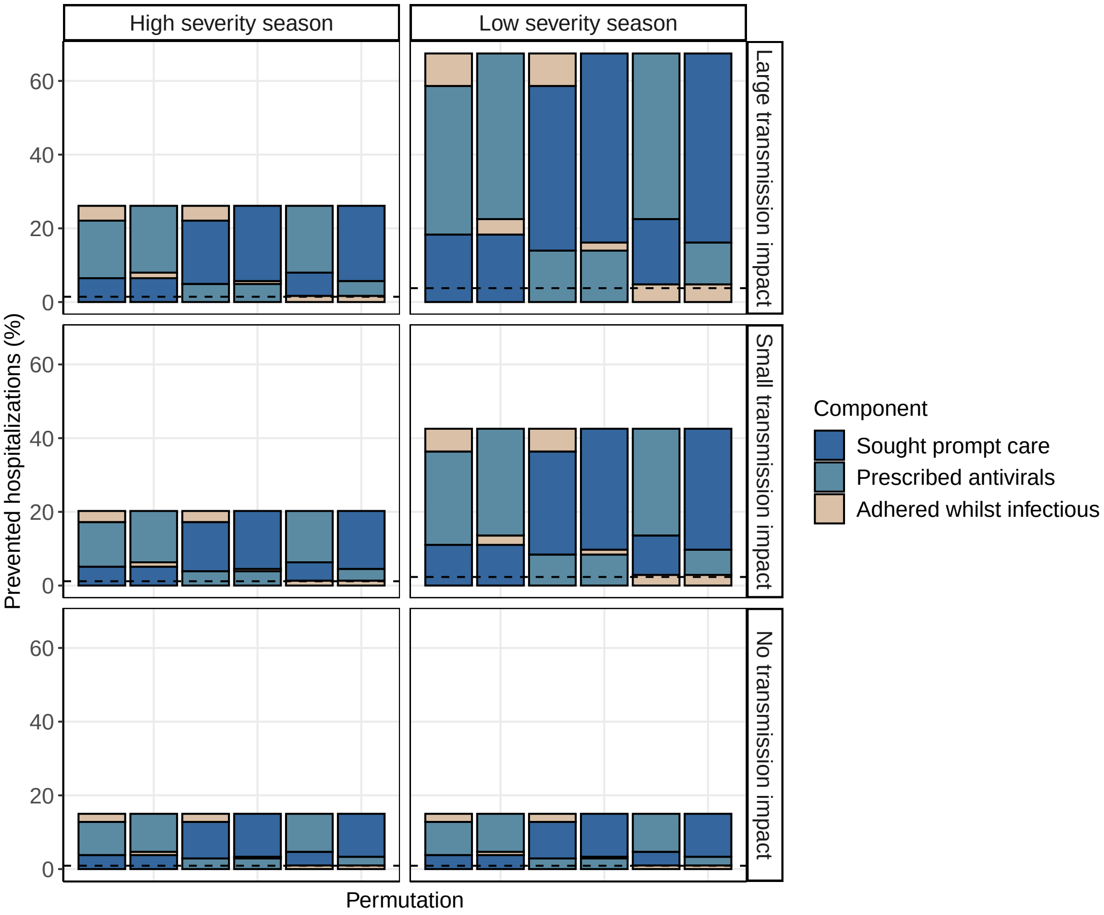


**Figure S9 – Impact of increasing all components that determine the overall percentage of symptomatic individuals undergoing prompt influenza antiviral treatment to 100%.** Permutation refers to the different possible orders in which components can be increased; colors at the base of each bar indicate the component that was increased first, colors in the middle indicate those that were increased second, etc. Bars represent the total percent of influenza-associated hospitalizations prevented in each antiviral effectiveness scenario relative to baseline. The dashed horizontal line represents the percent of hospitalizations prevented when all components are fixed at their mean starting values. 'Prompt care' refers to care sought within 48 hours of symptom onset.

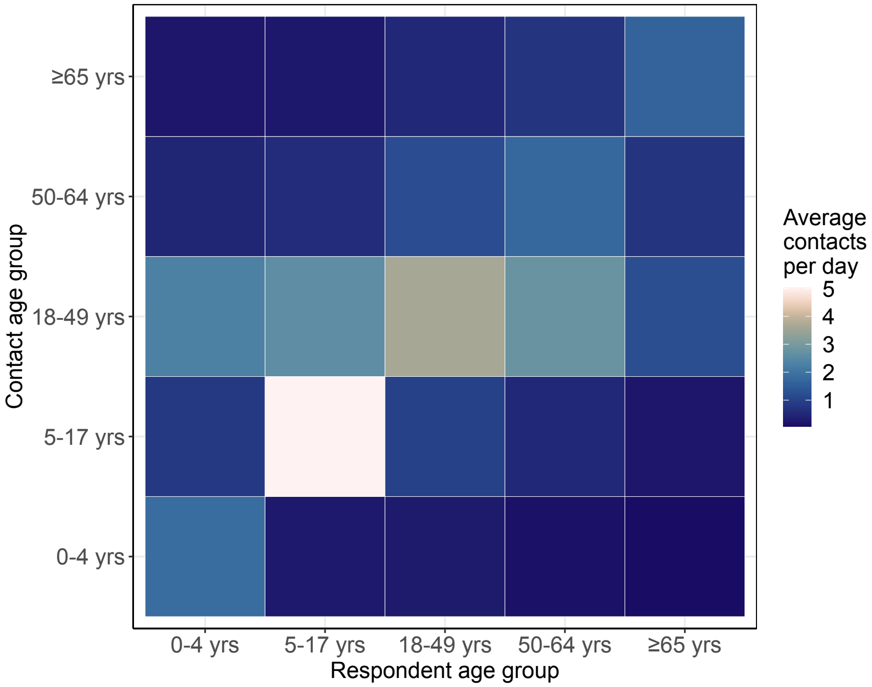


**Figure S10 – Contact matrix, $\hat{C}$.**

[16] Tsang TK, Wang C, Fang VJ, et al. Reconstructing household transmission dynamics to estimate the infectiousness of asymptomatic influenza virus infections. Proc Natl Acad Sci U S A. 120(33):e2304750120. doi:10.1073/pnas.2304750120
